# Deep learning-based Segmentation of Multi-site Disease in Ovarian Cancer

**DOI:** 10.1101/2023.01.10.22279679

**Authors:** Thomas Buddenkotte, Leonardo Rundo, Ramona Woitek, Lorena Escudero Sanchez, Lucian Beer, Mireia Crispin-Ortuzar, Christian Etmann, Subhadip Mukherjee, Vlad Bura, Cathal McCague, Hilal Sahin, Roxana Pintican, Marta Zerunian, Iris Allajbeu, Naveena Singh, Sahdev Anju, Laura Havrilesky, David E. Cohn, Nicholas W. Bateman, Thomas P. Conrads, Kathleen M. Darcy, G. Larry Maxwell, John B. Freymann, Ozan Öktem, James D. Brenton, Evis Sala, Carola-Bibiane Schönlieb

## Abstract

**Purpose:** To determine if pelvic/ovarian and omental lesions of ovarian cancer can be reliably segmented on computed tomography (CT) using fully automated deep learning-based methods.

**Materials and Methods:** A deep learning model for the two most common disease sites of high grade serous ovarian cancer lesions (pelvis/ovaries and omentum) was developed and compared against the well-established “no-new-Net” (nnU-Net) framework and unrevised trainee radiologist segmentations. A total of 451 pre-treatment and post neoadjuvant chemotherapy (NACT) CT scans collected from four different institutions were used for training (n=276), hyper-parameter tuning (n=104) and testing (n=71) of the methods. The performance was evaluated using the Dice similarity coefficient (DSC) and compared using a Wilcoxon test on paired results

**Results:** Our model outperforms the nnU-Net framework by a significant margin for both disease (validation: p=1×10-4,1.5×10-6, test: p=0.004, 0.005) and it does not perform significantly different from a trainee radiologist for the pelvic/ovarian lesions (p=0.392). On an independent test set (n=71), the model achieves a performance of 72±19 mean DSC for the pelvic/ovarian and 64±24 for the omental lesions.

**Conclusion:** Automated ovarian cancer segmentation on CT using deep neural networks is feasible and achieves performance close to a trainee-level radiologist for pelvic/ovarian lesions.

**Summary:** Deep learning-based models were used to assess whether fully automated segmentation is feasible for the main two disease sites in high grade serous ovarian cancer.

**Key Points:**

- First automated approach for pelvic/ovarian and omental ovarian cancer lesion segmentation on CT images.
- Automated segmentation of ovarian cancer lesions can be comparable with manual segmentation of trainee radiologists with three years of experience in oncological and gynecological imaging.
- Careful hyper-parameter tuning can provide models significantly outperforming strong state-of-the-art baselines.

## Introduction

After decades of unchanged treatment regimens for patient with ovarian cancer and little improvement in patients’ survival, this treatment landscape is currently changing, and an increasing number of therapeutic options can be offered. For standard treatments with chemotherapy, the interpretation of oncological computed tomography (CT) scans by an expert radiologist for evaluation of tumor spread usually includes only a small number of uni- or bidimensional lesion measurements that typically follow response evaluation criteria in solid tumors (RECIST 1.1) guidelines (1). However, these measurements are subjective, lack sensitivity for the early detection of treatment response and progression (2), and show only limited correlation with patient outcome (3,4). Novel treatments like immunotherapy, for example, require dedicated guidelines (5) and often several months of treatment monitoring before pseudo progression can reliably be distinguished from response. The development of non-invasive imaging biomarkers for response assessment as well as patient selection for such treatments is still in its infancy (6). Detailed volumetric response assessment as well as radiomics, have the potential to improve clinical decision making for both, patients undergoing standard of care chemotherapy and novel treatments. However, both require manual segmentation of the entire tumor burden by a radiologist. In advanced stage ovarian cancer with multi-site peritoneal disease, the detailed segmentation and annotation of a single scan can become a very time-consuming task and is only done for research purposes thus omitting potentially relevant information from clinical decision making.

Recently, deep neural networks based on the U-Net architecture (7) have shown promising results in challenging medical image segmentation problems. For example, the nnU-Net framework (8,9) achieved state-of-the-art performance in various biomedical segmentation challenges, such as the Medical Segmentation Decathlon (10). A recent survey also showed that nine out of ten top two performing teams in the 2020 MICCAI segmentation challenges are built using the nnU-Net framework as a baseline (11).

Deep neural networks are a promising solution for time-efficient and observer-independent segmentation of ovarian cancer lesions. Such methods allow volumetric response assessment of the disease instead of the currently used RECIST guidelines (1) and have the potential to reduce the manual annotation time (12,13) allowing researchers to create large scale datasets with high quality segmentations. Such datasets enable the creation of future disease quantification and response prediction tools to support clinical decision making and facilitate the development of clinical trials.

This is the first paper to propose a deep learning (DL) based approach for the automated segmentation of the disease located in the pelvis/ovaries and omentum, which are the predominant disease sites. A recently proposed automated approach based on classical machine learning focused on perihepatic and perisplenic ovarian cancer metastases segmentation (14,15) but did not address the more common locations.

## Materials and Methods

### Datasets

Patients were recruited prospectively into the respective studies. All images used in this study were acquired per clinical request and subsequently collected after informed patient consent was obtained for use in research approved by the local ethical review board. We retrospectively collected scans using only contrast enhanced axial CT images from patients with high grade serous ovarian carcinoma (HGSOC). The diagnosis of HGSOC was confirmed in all patients through biopsy and histopathological analysis. Patients without contrast enhanced CT scans or with unclear histopathological diagnosis were excluded.

For this study, we had a total of n=451 scans from four institutions and two countries available. As the majority (n=380) of data was obtained in the UK, we decided to use this part of the data for training and validation of the method. In particular, the larger subset (n=276) obtained at Addenbrooks Hospital Cambridge was used for training and the remaining scans (n=104) from St. Bartholomews Hospital London were used for validation. The remaining scans (n=71) obtained in the Gynecologic Cancer Center of Excellence program and the Cancer Imaging Archive (https://www.cancerimagingarchive.net/) from the US were used as a test set.

The patients across all datasets were treated with either immediate primary surgery (IPS) or three to six cycles of NACT. Among the 157 patients in the training dataset, 119 were treated with NACT for which both pre- and post-treatment scans were available. The remaining 38 patients were treated with IPS for which only the pre-treatment scan was available. The scans in the validation set were obtained from 53 patients that were treated with NACT. For all patients both pre- and post-NACT scans were available. However, two post-NACT scans were removed from the dataset as no disease was visible anymore after the treatment. All patients in the test data received IPS. Only pre-treatment scans are contained in this dataset.

The dataset compositions, including information on the patient age and quantitative measures on the two disease sites are shown in Table 1 and further details describing the heterogeneity of the acquisition protocols are provided in the Supplementary Materials. Further information on the patients such as ethnicity and clinical condition were not available to us due to the anonymization of the scans. Examples of the two disease sites are shown in Figure 1.

**Table 1.** Composition of the three datasets (total number of scans: 451) including information on voxel spacing and disease expression along all available time points. Due to the anonymization of the datasets, no information on the ethnicity of the patients was prevalent and the patient age was not available in 14 out of 71 patients in the test data. Additional information on the acquisition settings of the Computed Tomography scanners is given in the Supplementary Materials. NACT = neoadjuvant chemotherapy

| Dataset | Training | Validation | Test |
| --- | --- | --- | --- |
| Number of scans | 276 | 104 | 71 |
| Pre-treatment scans | 157 | 53 | 71 |
| Post-NACT scans | 119 | 51 | 0 |
| <u>Patient age [years]</u> |  |  |  |
| median | 65.5 | 65.5 | 63 |
| min-max | 29-90 | 35-85 | 41-80 |
| <u>Pixel spacing [mm]</u> |  |  |  |
| median | 0.68 | 0.76 | 0.77 |
| min-max | 0.53-0.93 | 0.61-0.96 | 0.57-0.98 |
| <u>Slice thickness [mm]</u> |  |  |  |
| median | 5.0 | 5.0 | 5.0 |
| min-max | 1.25-5.0 | 1.5-5.0 | 2.0-7.5 |
| <u>Pelvic/ovarian tumor</u> |  |  |  |
| Number of scans showing tumor in this location | 246 | 102 | 69 |
| Mean volume [cm<sup>3</sup>] | 275 | 241 | 381 |
| Mean number of connected components | 2.4 | 2.6 | 2.4 |
| <u>Omental tumor</u> |  |  |  |
| Number of scans showing tumor in this location | 198 | 98 | 56 |
| Mean volume [cm<sup>3</sup>] | 119 | 197 | 146 |
| Mean number of connected components | 6.7 | 5.3 | 5.7 |

**Figure 1.**
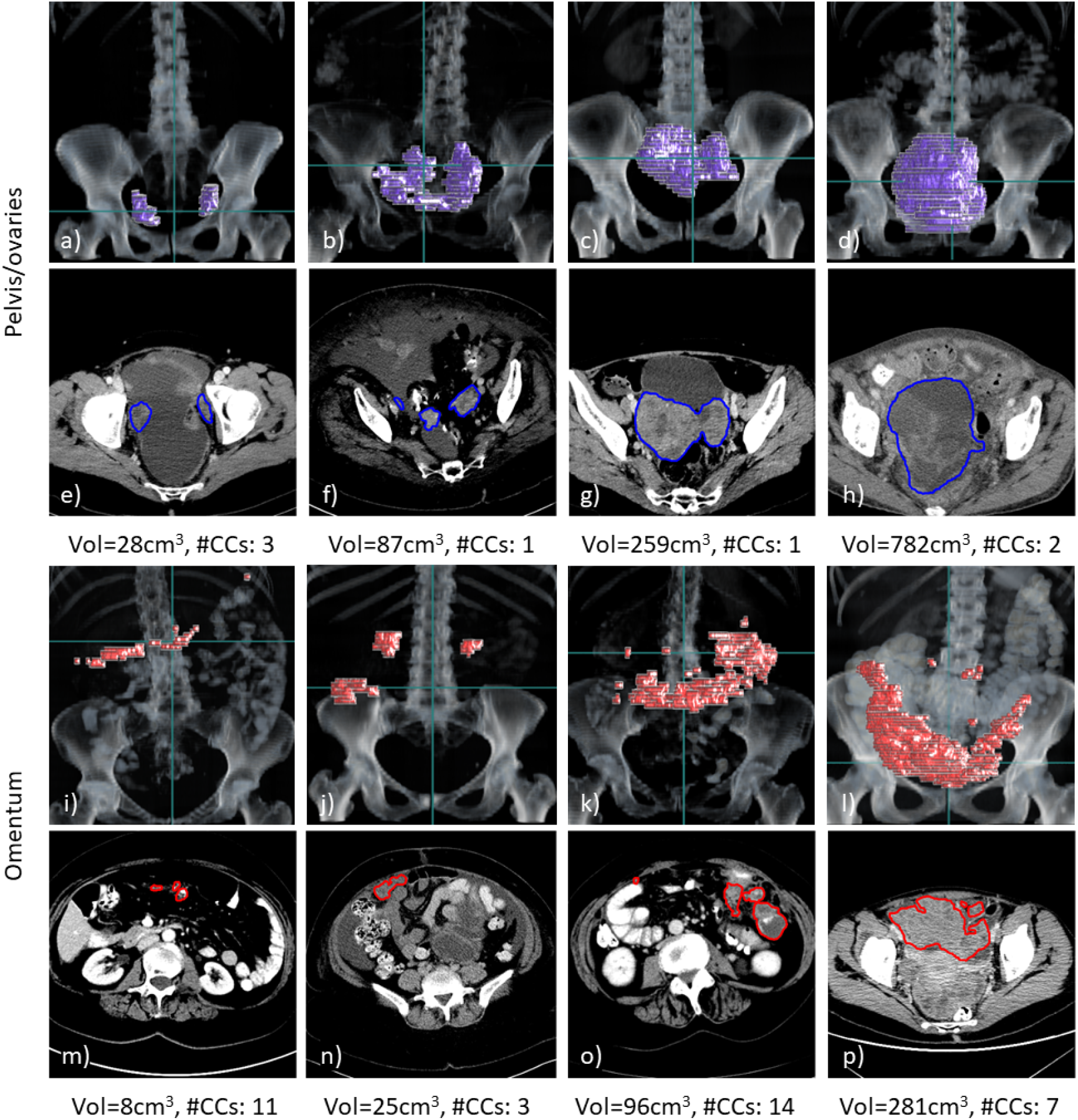
Examples of 3D volume renderings (a-d, i-l) and axial slices (e-h, m-p) for pelvic/ovarian and omental lesions of HGSOC patients. For each example, the ground truth tumor volume (Vol) and number of connected components (#CCs) are shown. The scans shown were all contained in the training set and selected such that their lesion volume equals the 25, 50, 75 and 90 percentiles of the lesion volume in the training set (left to right). The horizontal green line in the rendering visualizations correspond to the axial slice shown below. Both disease sites demonstrate a great variability of disease expression among different patients, which poses a great challenge for manual and automated segmentation methods.

### Manual Annotation

All manual segmentations were performed using Microsoft Radiomics application (project InnerEye; https://www.microsoft.com/en-us/research/project/medical-image-analysis/), Microsoft, Redmond, WA, USA).

All segmentations used as ground truth in this study were either manually segmented or corrected by RW (consultant radiologist with ten years of experience in oncological and gynecological imaging). The training data set was segmented solely by RW. The validation data set was pre-segmented by a trainee radiologist (three years of experience in oncological and gynecological imaging) and subsequently reviewed and corrected by RW. For all scans in this dataset, both the unrevised trainee and the ground truth segmentations were available. The test set was segmented by HS (initials blinded; six years of experience in oncological and gynecological imaging) and reviewed by RW. All segmentations in this dataset were found to be of satisfying quality; therefore, only one set of segmentations (ground truth) was available for these scans.

### Deep Learning Model

As no literature exists on the segmentation of pelvic/ovarian and omental lesions in ovarian cancer, we first used the 3D full resolution U-Net of the nnU-Net framework (8,9) as a baseline for this segmentation task. The framework automatically adapts to new datasets, suggests model hyper-parameters and can be considered the current state-of-the-art in 3D biomedical image segmentation (10, 11). Our model was obtained by reimplementing the nnU-Net framework from scratch, benchmarking both implementations and performing extensive hyper-parameter optimization. As already observed by the authors of nnU-Net (8,9), we did not find an impact of minor architectural changes to the performance of our model. Instead, we decided to focus on fine tuning the hyper-parameters suggested by the nnU-Net framework.

As all current state-of-the-art 3D biomedical segmentation networks, nnU-Net is based upon the U-Net architecture (7,11). For our dataset nnU-Net suggested a simple U-Net with six stages, LReLU activation functions, Instance Normalization and 32 filters in the first block doubling at each stage. We found it beneficial to reduce the number of stages to four and replace the encoder with a ResNet (16) of 1, 2, 6, and 3 blocks per stage. The nnU-Net framework further suggests training the networks for 250.000 batches of size 2 using an SGD optimizer with Nesterov’s momentum of factor 0.99, weight decay of 3×10-5, and a polynomial decaying learning rate from 0.01 to 0. We instead found it beneficial to increase the batch size to 4, the weight decay to 10-4, decrease the momentum to 0.98 and change the learning rate schedule to a linear ascent plus cosine decay with maximum learning rate 0.02.

The framework uses resizing of voxel spacing and Z-normalization of the grey values as preprocessing and various spatial (rotation, scaling, flipping) and grey value based (Gaussian noise, multiplicative scaling, contrast, blurring, gamma) transformations on the fly for data augmentation. During inference, a Sliding Window algorithm with a Gaussian weighting and test-time augmentations by evaluating all eight permutations of flipping over x-, y- and z-axis was applied. We found no benefit in changing the pre-processing, data augmentation and evaluation based on the Sliding Window algorithm, hence they were left unchanged and applied as suggested by the authors of nnU-Net (8,9).

### Code and Model Availability

All code was developed using Python (version 3.9.4) as a programming language and the deep learning framework PyTorch (version 1.9.0). To make the training reproducible and share our model with the research community, we made the training code, inference code and model hyper-parameters and weights publicly available on our code repository at https://github.com/ThomasBudd/ovseg.

### Statistical Analysis

To test whether differences in DSC were significant, we computed *p*-values using the Wilcoxon test on paired results. These computations were performed using the Python package SciPy.

## Results

### Performance assessment

All metrics were only computed on scans containing the corresponding disease site. The results (expressed as mean ± standard deviation) are summarized in Figure 2 and Supplementary Table 1. On the validation set our model achieved mean DSCs of 66 and 51 for pelvic/ovarian and omental lesions respectively outperforming the baseline by a significant margin (p=1x and 1.5x). On the test set our model achieved mean DSCs of 72 and 64 which was also found to be a significant improvement over the baseline (p=0.004 and 0.005). Additionally, we compared the DSCs achieved by both automated methods and the trainee radiologist on the validation set. Both models performed significantly worse than the trainee radiologist for the segmentation of omental lesions (p=1x and 3.6x). However, considering the pelvic/ovarian lesions, the baseline model performs significantly inferior to the trainee radiologist (p=0.041), while there was no such significant difference between our proposed model and the trainee radiologist (p=0.392).

**Figure 2.**
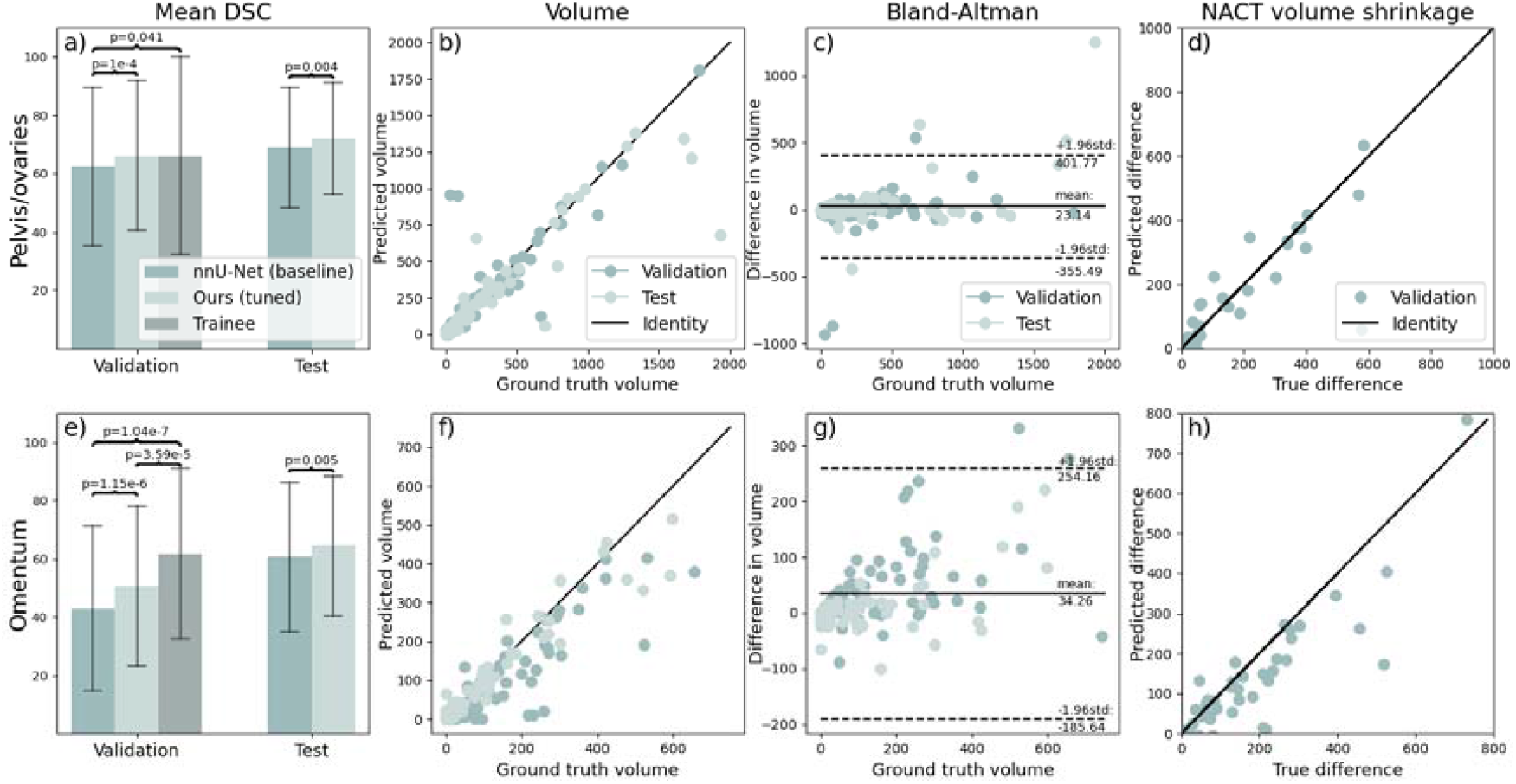
Evaluation of model performance on unseen datasets in terms of DSC (a,e) and volume (b-d, f-h). Trainee radiologist segmentations were only available on the validation set. The brackets indicate significant differences. All volumes are given in cm^3^. It can be observed in panels a) and e) that our method outperforms the nnU-Net baseline for both sites on the validation and test set and that our method does not perform significantly different from a trainee radiologist in segmenting pelvic/ovarian lesions in contrast to nnU-Net. Panels b-d and f-h suggest that the model in its current state can be used to determine disease volume for both sites. DSC = Dice similarity coefficient.

Additionally, we compared the disease volume of the ground truth with the automated annotations of our proposed model. Despite the presence of some outliers, Figure 2 shows close agreement in volume for both disease sites considering the volume comparison of individual scans (second column) and the difference of pre- and post-NACT volume. The Bland-Altman plot shows that on average the ground truth volume is greater than the predicted volume for both sites.

Examples of automated and trainee radiologist segmentations are presented in the left and middle columns of Figure 3.

**Figure 3.**
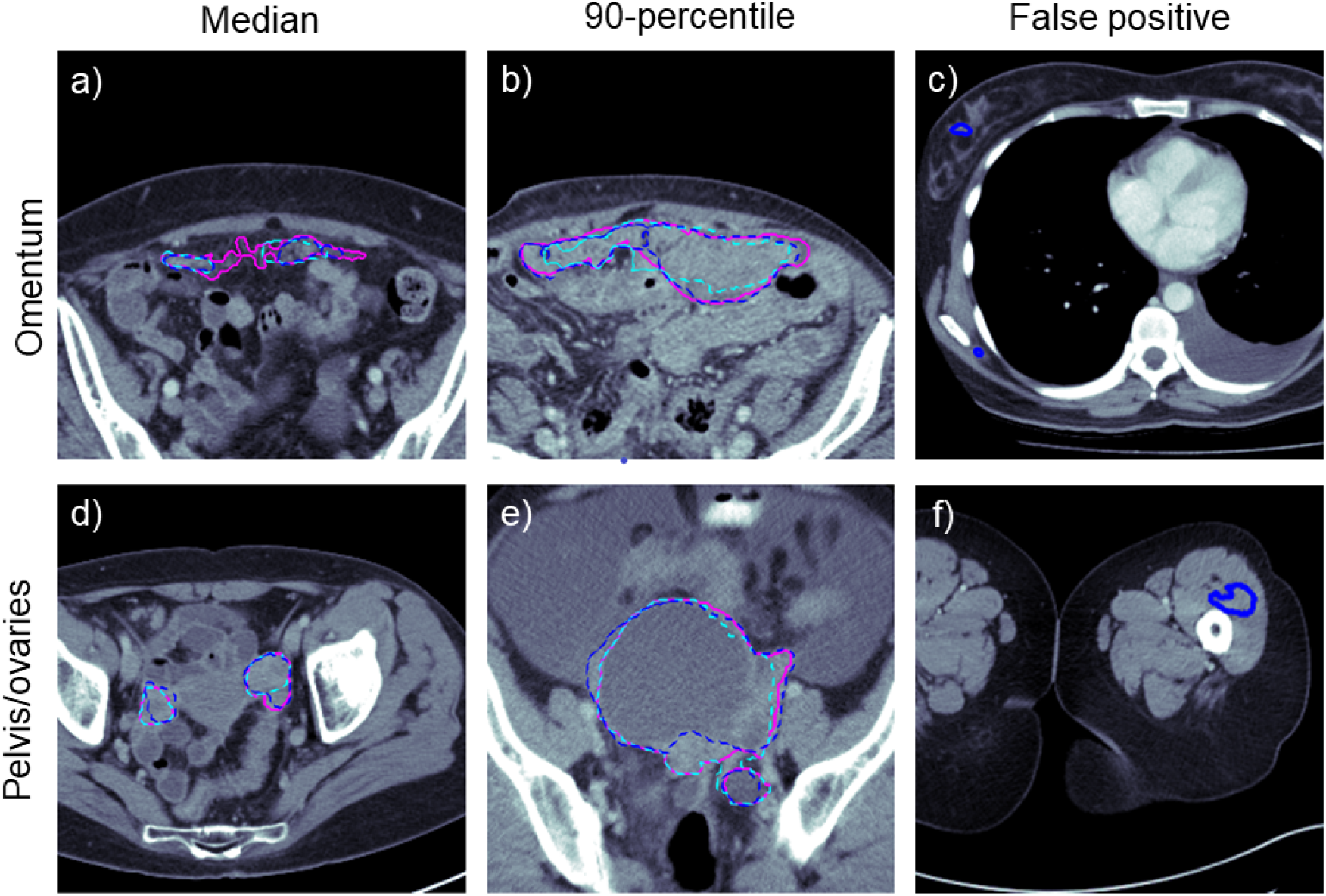
Examples of ground truth, automated and trainee radiologist segmentations (pink, cyan and blue respectively). The first two columns (a, b, d, e) show the cases with median and 90-percentile DSC from the pooled validation and test set. The visual comparison between the automatically generated and manual trainee radiologist segmentation demonstrates typical mistakes of the two instances. Both seem to struggle with the inclusion and exclusion of objects close to the segmentation boundary. The last column (c, f) shows examples of outliers at the extreme ends of the volumes. The segmentation model confused dense components of breast tissue with omental disease as both are embedded in fat.

### Outlier and Error Analysis

To assess common errors and outliers, the validation and test set were pooled (n=175) and inspected visually and quantitatively.

Low DSC values were regularly found on scans with low ground truth volume. In the subset of scans with the bottom 25% DSC performance, lower median disease volume was found compared to the full set (omentum: 6 vs. 21 cm^3^, pelvis/ovaries: 9 vs. 37 cm^3^). Vice versa, on the scans with the 25% lowest volumes, lower mean DSC performance was found when compared to the full set (omentum: 35 vs. 55, pelvis/ovaries: 46 vs. 68).

A visual evaluation revealed common false positive predictions in the extreme ends of the scan volumes outside of anatomic regions where disease commonly occurs. On 41 scans omental disease was falsely annotated in breast tissue, in 13 cases near the scapulae, and in five cases false positive annotations were found in the thighs. Examples of such false positive annotations can be found in the right column of Figure 3.

To better understand the influence of false positive and false negative predictions we computed the sensitivity and precision. For both disease sites the sensitivity was found to be lower than the precision (omentum: 52 vs. 70, pelvis/ovaries: 69 vs. 73).

Another source of error was the confusion between classes. We found that in 12 out of 170 cases containing pelvic/ovarian disease, at least parts of the ground truth annotation intersected with automated annotation of omental disease. Vice versa in 18 out of 155 scans containing omental disease at least one voxel of omental disease was marked as pelvic/ovarian disease by the algorithm. Even to the trained radiologist’s eye, it can be challenging to distinguish between extensive pelvic and omental disease when tumors form conglomerates.

## Discussion

This work presents the first automated deep learning-based ovarian cancer CT segmentation approach for the two main disease sites: the pelvis/ovaries and the omentum, while previous work only addressed less common disease sites (14,15). While the relatively low DSC values suggest inferior performance compared to expert consultant radiologists, we could demonstrate similar performance to a trainee radiologist with three years of experience for the pelvic/ovarian lesions despite the complexity of the disease and using only a few hundred scans for training. Further, we demonstrated that our model significantly outperforms the well-established state-of-the-art framework nnU-Net and generalizes to a test set from a different country. The heterogeneity of the disease between patients was demonstrated in Figure 1. This causes great challenges for both manual and automated segmentation and might be the main reason for the performance gap between our model and the ground truth. The fact that our model performed better on the test set compared to the validation set, for which the hyper-parameters were tuned, is most likely a result of heterogeneity between the datasets.

We believe that the model in its current form is already of clinical relevance. Previous approaches have already demonstrated that deployed deep learning models can decrease the manual preparation time in clinical routine (12) and in a research setting for the creation of large scale datasets (13). These datasets might ultimately allow the creation of sophisticated chemotherapy response or survival prediction models (17) and improve patient care. Further, Figure 2 suggests that the model might be ready to allow volumetric assessment of the disease without requiring manual interventions. This might be of particular interest for centers without specialization in high grade serous ovarian cancer.

Our main limitations are the following. The imbalance between sensitivity and precision, which was especially large for the omental lesions, indicates that the DSC can be further improved by careful calibration of the model parameters as suggested in (18). This might also be a solution for underestimation of disease volume as shown in the Bland-Altman plots of Figure 2. The false positive predictions in the extreme ends of the scans, such as the breast tissue and the lower limbs, might be removed in two different ways. Firstly, organ segmentation (13) can be applied to identify the regions in which the lesions typically occur. Secondly, the patch sampling scheme can be modified as the currently employed schemes undersample the extreme ends of the volumes. Next, augmenting the model with other frameworks can be attempted to improve the performance. For example, a recently introduced vision transformer-based framework has shown inferior performance when being compared to nnU-Net but could demonstrate the ensembling both methods outperform the standalone frameworks (19). In addition, future approaches should integrate the other disease sites of HGSOC into automated segmentation approaches. However, those occur less frequently, are on average of lower volume and often spread throughout the whole abdomen and beyond, thus imposing challenges for automated segmentation models. For the training of future models, it is also desirable to have access to larger datasets with high quality annotations. This was not available to us in this initial study as annotations are time-consuming to obtain and experts for this disease are rare. We believe that larger datasets along with continuously exploring new training methods will help closing the performance gap between the consultant radiologists and the deep learning model.

To summarize, we presented the first deep learning-based approach for ovarian cancer segmentation on CT and the first automated approach for the segmentation of pelvic/ovarian and omental lesions and demonstrated both state-of-the-art performance, as well as common errors of our method.

## Data Availability

All CT scans and segmentations used for training, validation and testing of the methods are not publicly available, except for the 20 scans obtained from the Cancer Imaging Archive.
The trained models are publicly available.

https://github.com/ThomasBudd/ovseg

## Abbreviations

3D: three-dimensional
CT: Computed Tomography
DSC: Dice similarity coefficient
HGSOC: high grade serous ovarian carcinoma
IPS: immediate primary surgery
NACT: neoadjuvant chemotherapy
nnU-Net: no-new-Net

## Acknowledgments

The authors would like to thank Fabian Isensee for both making the nnU-Net library freely available and easy to use for new datasets.

This work was partially supported by The Mark Foundation for Cancer Research and Cancer Research UK Cambridge Centre [C9685/A25177], the Wellcome Trust Innovator Award [RG98755] and the CRUK National Cancer Imaging Translational Accelerator (NCITA) [C42780/A27066].

Additional support was also provided by the National Institute of Health Research (NIHR) Cambridge Biomedical Research Centre (BRC-1215-20014). The views expressed are those of the authors and not necessarily those of the NHS, the NIHR, or the Department of Health and Social Care.

This project has been funded in whole or in part with Federal funds from the National Cancer institute, National Institutes of Health, Department of Health and Human Services, under Contract No. 75N91019D00024.

CBS acknowledges support from the Leverhulme Trust project on ‘Breaking the non-convexity barrier’, the Philip Leverhulme Prize, the Royal Society Wolfson Fellowship, the EPSRC grants EP/S026045/1 and EP/T003553/1, EP/N014588/1, EP/T017961/1, European Union Horizon 2020 research and innovation programmes under the Marie Skodowska-Curie grant agreement No. 777826 NoMADS and No. 691070 CHiPS, the Cantab Capital Institute for the Mathematics of Information and the Alan Turing Institute.

The work by Öktem was supported by the Swedish Foundation of Strategic Research under Grants AM13-0049.

Microsoft Radiomics was provided to the Addenbrooke’s Hospital (Cambridge University Hospitals NHS Foundation Trust, Cambridge, UK) by the Microsoft InnerEye project.

## Supplementary Materials

### Acquisition Characteristics of the Analyzed Ovarian Cancer CT Datasets

There is high variation in image acquisition parameters and scanner manufacturers across all three datasets. In the training data, the most used scanner manufacturer was Siemens followed by GE, Philips, and Toshiba with 204, 56, 12 and 4 scans, respectively. In this dataset, one, 45, 115, 110 and 5 scans were acquired at a KVP of 80, 100, 120, 130 and 140 respectively. In the validation data, the most used scanner manufacturer was Siemens followed by Toshiba, Philips, and GE with 64, 24, 8 and 8 scans, respectively. Here all scans were acquired at a KVP of 120, except one for which 100 KVP was used. In the test dataset, 38 scans were acquired using GE scanners, followed by 26 Siemens, four Toshiba, one Philips, one Imatron, and one MPTronic scanner. In this dataset eight, 58, and five scans were acquired at 100, 120, 130 KVP respectively.

We analyzed the influence of the acquisition protocol on the performance of the model in terms of DSC. Therefore, we pooled the validation and test set. As the resulting dataset only showed large heterogeneity in terms of manufacturer, but not in terms of KVP we decided to focus only on the influence of the manufacturer to the performance and removed the two scans acquired my Imatron and MPTronic scanners from the dataset due to low sample size. Supplementary Figure 1 shows the manufacturer of the CT scanner versus the performance of the model in terms of DSC. It can be observed that differences are difficult to spot by eye due to the high variability of the DSC. We further compared each manufacturer against the remaining ones using the Wilcoxon Rank test. For the pelvic/ovarian disease we computed the p-values 0.296 (GE Medical Systems), 0.527 (Siemens), 0.743 (Toshiba), and 0.244 (Philips). For omental disease the p-values were computed as 0.625 (GE Medical Systems), 0.292 (Siemens), 0.043 (Toshiba), and 0.887 (Philips).

To summarize, in all except one case no significantly different performance in dependence of the scanner manufacturer could be found.

### Hyper-parameter tuning

In the first step of our hyper-parameter tuning we reduced the in-plane resolution of the scans from 0.67 mm (as suggested by nnU-Net) to 0.8mm, which later allowed us to reduce the patch size. We also applied progressive learning by splitting the training into four quarters and reducing both the number of voxels per sample and the magnitude of the grey value augmentations by a factor of four, three and two over the first three quarters of the training. The last quarter was applied as usual.

Next, we applied several architectural modifications, but only found increase the capacity of the network to by replacing the decoder with a ResNet to be beneficial. In detail we changed the six stage U-Net (i.e. a U-Net with five downsampling operations in the decoder) to a four stage U-Net with 1, 2, 6 and 3 residual blocks in the corresponding stages. We used standard residual blocks where each block consisted of two convolution-Instance normalization-LReLU units followed by a skip connection. As now the number of downsampling operations was reduced from five to three we could reduce the patch size from 224 to 216 which reduced the training time again.

Lastly, we found a great improvement by altering the convergence parameters. We found a benefit in changing the learning rate schedule from the almost linear decay from 0.01 to 0 as suggested by nnU-Net to a linear ascent plus cosine decay with maximum learning rate 0.02.

The increase in maximum learning rate was not possible without using the linear warmup, which happened over the first five percent of the training. Next, we made use of the full 24GB of VRAM in our GPUs and increased the batch size from two to four. Initially this caused a performance decrease, but we finally obtained an increase in performance when decreasing the momentum factor of the SGD from 0.99 to 0.98 and increasing the weight decay from 3×10-5 to 10-4.

**Supplementary Table 1.** Model and trainee performance on unseen datasets in terms of DSC (mean ± std). Significant differences between our model and the baseline and the trainee and our model are marked with an asterisk. Trainee radiologist segmentations were only available on the validation set. Our implementation is available at https://github.com/ThomasBudd/ovseg.

|  | Implementation | Validation |  | Test |  |
| --- | --- | --- | --- | --- | --- |
|  |  | pelvis/ovaries | omentum | pelvis/ovaries | omentum |
| Baseline | nnU-Net (6,7) | 62±27 | 43±28 | 69±21 | 60±26 |
| Tuned | Ours | 66±26* | 51±27* | 72±19* | 64±24* |
| Trainee | | 66±34 | 62±29* | - | - |

**Supplementary Figure 1.**
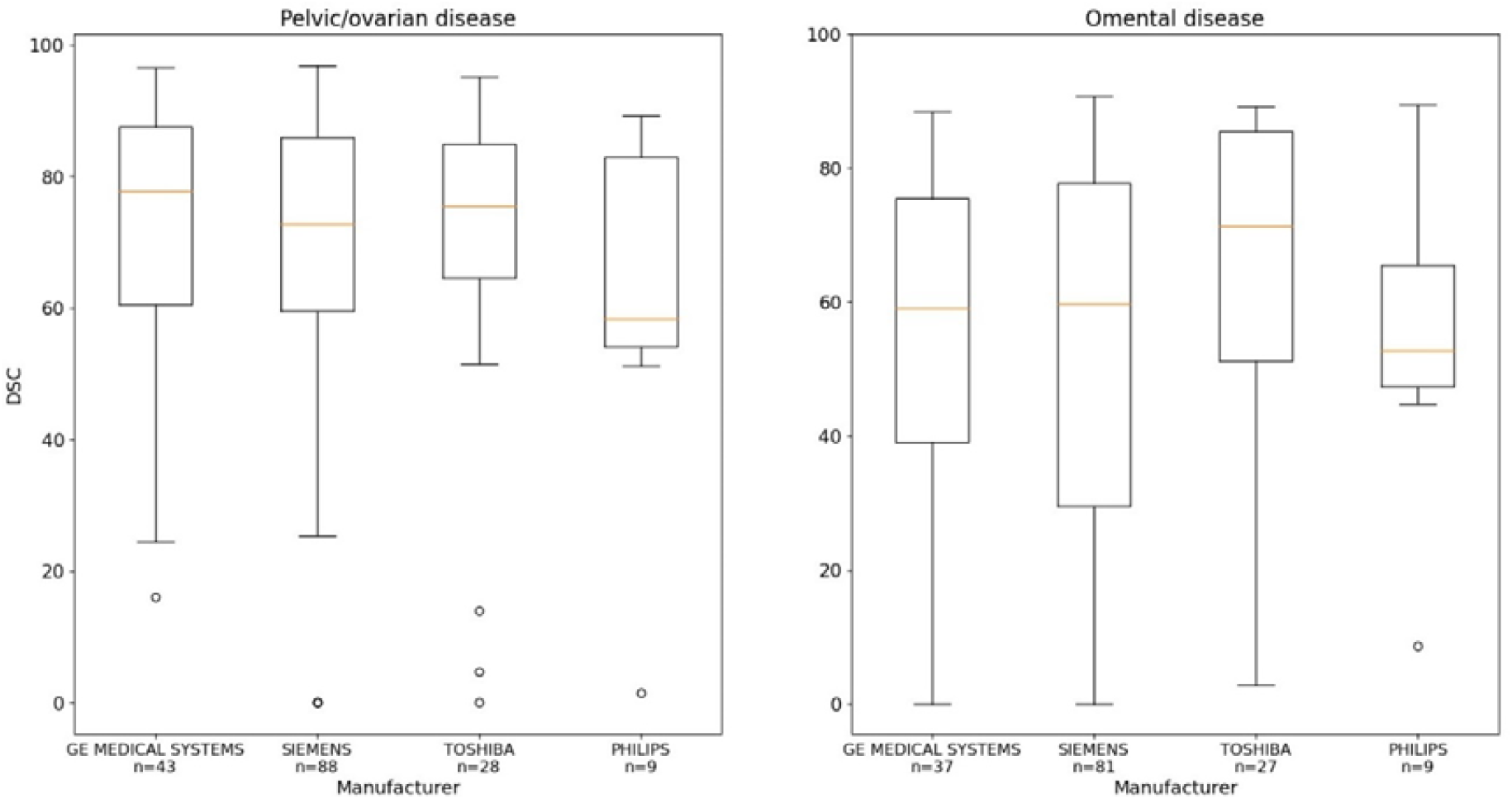
Comparison of the models performance in terms of DSC on scanners from different manufacturers. DSC = Dice similarity coefficient

