## Supplementary Materials for "Deep learning-based Segmentation of Multi-site Disease in Ovarian Cancer"

**Acquisition Characteristics of the Analyzed Ovarian Cancer CT Datasets**

There is high variation in image acquisition parameters and scanner manufacturers across all three datasets. In the training data, the most used scanner manufacturer was Siemens followed by GE, Philips, and Toshiba with 204, 56, 12 and 4 scans, respectively. In this dataset, one, 45, 115, 110 and 5 scans were acquired at a KVP of 80, 100, 120, 130 and 140 respectively. In the validation data, the most used scanner manufacturer was Siemens followed by Toshiba, Philips, and GE with 64, 24, 8 and 8 scans, respectively. Here all scans were acquired at a KVP of 120, except one for which 100 KVP was used. In the test dataset, 38 scans were acquired using GE scanners, followed by 26 Siemens, four Toshiba, one Philips, one Imatron, and one MPTronic scanner. In this dataset eight, 58, and five scans were acquired at 100, 120, 130 KVP respectively.

We analyzed the influence of the acquisition protocol on the performance of the model in terms of DSC. Therefore, we pooled the validation and test set. As the resulting dataset only showed large heterogeneity in terms of manufacturer, but not in terms of KVP we decided to focus only on the influence of the manufacturer to the performance and removed the two scans acquired my Imatron and MPTronic scanners from the dataset due to low sample size. Supplementary Figure 1 shows the manufacturer of the CT scanner versus the performance of the model in terms of DSC. It can be observed that differences are difficult to spot by eye due to the high variability of the DSC. We further compared each manufacturer against the remaining ones using the Wilcoxon Rank test. For the pelvic/ovarian disease we computed the p-values 0.296 (GE Medical Systems), 0.527 (Siemens), 0.743 (Toshiba), and 0.244 (Philips). For omental disease the p-values were computed as 0.625 (GE Medical Systems), 0.292 (Siemens), 0.043 (Toshiba), and 0.887 (Philips).

To summarize, in all except one case no significantly different performance in dependence of the scanner manufacturer could be found.

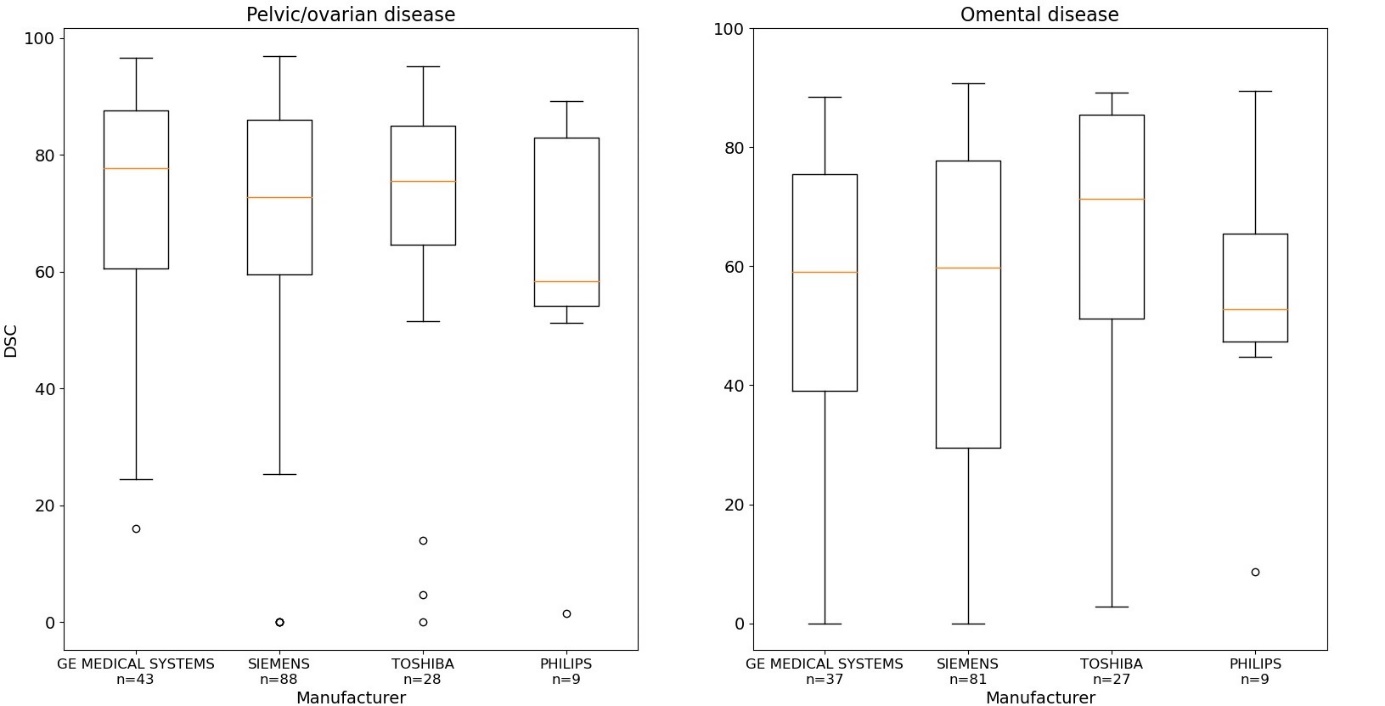

**Supplementary Figure 1.** Comparison of the models performance in terms of DSC on scanners from different manufacturers.

**Hyper-parameter tuning**

In the first step of our hyper-parameter tuning we reduced the in-plane resolution of the scans from 0.67 mm (as suggested by nnU-Net) to 0.8mm, which later allowed us to reduce the patch size. We also applied progressive learning by splitting the training into four quarters and reducing both the number of voxels per sample and the magnitude of the grey value augmentations by a factor of four, three and two over the first three quarters of the training. The last quarter was applied as usual.

Next, we applied several architectural modifications, but only found increase the capacity of the network to by replacing the decoder with a ResNet to be beneficial. In detail we changed the six stage U-Net (i.e. a U-Net with five downsampling operations in the decoder) to a four stage U-Net with 1, 2, 6 and 3 residual blocks in the corresponding stages. We used standard residual blocks where each block consisted of two convolution-Instance normalization-LReLU units followed by a skip connection. As now the number of downsampling operations was reduced from five to three we could reduce the patch size from 224 to 216 which reduced the training time again.

Lastly, we found a great improvement by altering the convergence parameters. We found a benefit in changing the learning rate schedule from the almost linear decay from 0.01 to 0 as suggested by nnU-Net to a linear ascent plus cosine decay with maximum learning rate 0.02. The increase in maximum learning rate was not possible without using the linear warmup, which happened over the first five percent of the training. Next, we made use of the full 24GB of VRAM in our GPUs and increased the batch size from two to four. Initially this caused a performance decrease, but we finally obtained an increase in performance when decreasing the momentum factor of the SGD from 0.99 to 0.98 and increasing the weight decay from 3x10-5 to 10-4.

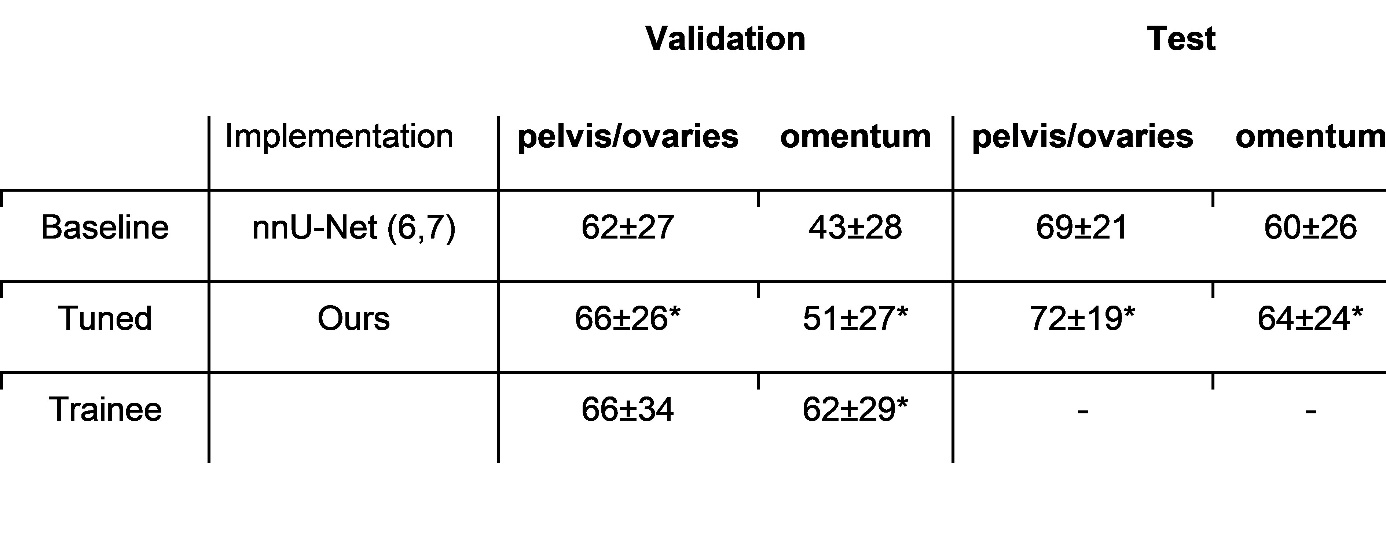

**Supplementary Table 1.** Model and trainee performance on unseen datasets in terms of DSC (mean ± std). Significant differences between our model and the baseline and the trainee and our model are marked with an asterisk. Trainee radiologist segmentations were only available on the validation set. Our models and implementation is available at https://github.com/ThomasBudd/ovseg.
